# Intervention provision and engagement in Colombia’s PAPSIVI – a national psychosocial support service for over half a million victims of armed conflict

**DOI:** 10.1101/2025.07.08.25331104

**Authors:** Charlotte Constable Fernandez, Alida Acosta-Ortiz, María Camila García Durán, Elisavet Pappa, Rob Saunders, Francesca Solmi, William Tamayo-Agudelo, Fabio Idrobo, Vaughan Bell

## Abstract

**Background:** Colombia’s PAPSIVI program is the world’s largest psychosocial support service for victims of armed conflict providing support for over half a million civilians. However, service delivery has only previously been examined in small studies, making it difficult to understand to what extent PAPSIVI delivers interventions that are adequately targeted to individuals with the most serious exposures to the armed conflict, and to what extent attendees remain engaged with interventions.

**Methods:** We investigated how different conflict exposures related to PAPSIVI intervention assignment and engagement. We linked anonymised national data from the register of victims of the Colombian armed conflict to data from N = 534,818 PAPSIVI attenders. Analysis used logistic and linear regression with cluster robust standard errors, adjusted for a range of potential confounders.

**Results:** Intervention types were broadly provided in line with PAPSIVI guidelines, with victims experiencing torture, sexual violence, and forced recruitment more likely to receive individual sessions, while community-level impacts received community interventions. Female sex, ethnic minority status, and receiving state-subsidised healthcare were associated with higher intervention engagement. Those with previous mental health diagnoses had increased odds of attending individual or family sessions but lower odds of group or community sessions, consistent with recommendations for more intensive intervention for those with higher mental health needs. 29% of individual session attendees only received a single session, potentially indicating early dropout or unsuitable service provision for a proportion of attendees.

**Conclusions:** This study provides insights into support provision for civilian victims of conflict indicating that psychosocial support provision can be managed effectively at very large scales.

## Background

The Colombian armed conflict has disproportionally affected civilians. A recent estimate from the Colombian Victim’s Unit reported that 18.3% of the population are recognised as registered victims of the conflict by the state [1] although this is likely to be an under-estimate given victim registration began in 1985, some 20 years after the recognised start of the conflict in 1964 [2]. The extended nature of the conflict has led to a high burden of poor mental health and damage to the social structure of communities [3]. Consequently, the provision of effective psychosocial support services for civilian victims of the conflict has been identified as a priority by both state policy and independent researchers [3–5]. A major challenge with providing adequate mental health and psychosocial care is the gap between the scale of need and the availability of services. Colombia has over 9.5 million officially registered victims of the conflict, including millions displaced by violence and hundreds of thousands subjected to widespread human rights abuses committed by all armed actors [6]. Concerns have been raised about the capacity of mainstream health services to provide adequate support, as they are disproportionately concentrated in urban centres and frequently distant from populations most affected by the conflict [7,8].

To address these challenges, the Colombian government developed a national service called PAPSIVI (an acronym from its Spanish-language title *Programa de Atención Psicosocial y Salud Integral a Víctimas*), which is the largest psychosocial and mental healthcare service yet developed for victims of armed conflict [9]. Therefore, evaluating PAPSIVI service provision is of international interest with implications for the effective design and implementation of support services for conflict-affected victims worldwide.

PAPSIVI’s psychosocial care programme is delivered by psychologists, social workers and community advocates. It is designed to address the psychosocial harm of the conflict on multiple levels – ranging from its impact on an individual’s mental health, to its effects on the family, and to damage to the social fabric of communities [10]. This multi-level approach is at the heart of PAPSIVI’s implementation and its psychosocial programme has been designed with four intervention modalities [11]: i) individual intervention involving counselling or psychotherapy, depending on the level of individual distress and disability; ii) small group interventions, focused on developing coping skills and sharing experiences between group members; iii) family interventions, aimed at helping improve family functioning after loss, displacement, or fragmentation, and iv) community level interventions, designed to enhance community cohesion through activities such as community problem-solving, strengthening cultural identity and belonging, and collective memorialisation.

Importantly, the PAPSIVI service documentation recommends certain intervention modalities for either specific psychosocial effects (e.g. guilt, mourning) or specific types of conflict exposure types (e.g. torture, kidnapping). These recommendations are based on the type, extent, and severity of conflict-related harm [10]. For example, individual and / or family sessions are recommended for sexual violence, forced disappearance, and forced recruitment to armed groups [12,13]. For individuals affected by forced recruitment, the guidelines suggest either individual, family or community sessions [12,14]. According to PAPSIVI intervention manuals, individual, group and family sessions are designed as eight session interventions, and community interventions as six session interventions. Practitioners are able to use their judgement to tailor intervention modality and duration for any specific case. However, it is not known the extent to which these exposure-based recommendations result in engagement with specific intervention modalities, as practitioners and service users may influence modality provision due to resources, personal preference, or professional judgment.

Successful adherence to service guidelines is considered a marker of service-delivery effectiveness [15,16] and a key component of establishing intervention fidelity [17]. Similarly, engagement with psychological intervention sessions is a widely-evidenced predictor of better outcome [18–20]. PAPSIVI was found to be acceptable with a high degree of attendee satisfaction in a mixed methods study including 771 nationally representative attenders [21]. However, no study to date has examined the extent to which different exposures to the armed conflict relate to intervention modalities and service engagement for victims of the Colombian armed conflict using the full national cohort.

To address this gap, the present study used anonymised data from Colombia’s register of victims of the armed conflict linked with PAPSIVI service data to understand to what extent different intervention modalities are being delivered to victims based on their exposures and to what extent attendees are remaining engaged with interventions. Specifically, this study aimed to: i) assess whether the type of intervention modalities (individual, group, community or family sessions) are differently provided depending on the type of victimisation experienced by individuals; ii) investigate whether individuals with pre-existing mental health diagnoses receive care aligned with PAPSIVI guidelines, namely, greater level of intervention intensity in terms of a greater number of sessions and individual interventions that are aimed at more serious presentations; and, ii) evaluate patterns of intervention engagement in terms of total sessions attended and the extent to which these differed by demographics reflecting membership of key marginalised groups.

## Methods

The project received ethical approval from both the Ethics Committee of the Fundación Santa Fe de Bogotá in Colombia (ID CCEI-15951-2023) and the University College London Research Ethics Committee in the United Kingdom (Project ID: 8275/002).

### Data sources

We used anonymised data from the central registry of victims of the Colombian armed conflict, known as the *Registro Unico de Víctimas* (RUV) in Spanish. This is a statutory list of conflict victims maintained by the Colombian government. Registration is legally required to access reparations and must be done at authorised offices, such as local government sites or the Victims’ Unit – the government agency responsible for registering victims of the armed conflict and delivering assistance. Applicants are interviewed about the time, place, and nature of the events and may need to provide supporting documents. Reparations available to victims are defined in law and depend on an assessment of the type and severity of harm and may include financial compensation, humanitarian aid, medical and psychosocial rehabilitation, land and property restitution. Victims are defined as having experienced victimisation personally or as a first-degree family member (parents, children and siblings) of someone affected by a victimising event. The types of victimising events ultimately recorded in the registry depend which events are declared by the applicant and the verification process. More than one victimising event can be recorded per individual although only one is needed to be recognised as an officially registered victim. The RUV data was linked with data indicating access to PAPSIVI via a shared pseudonymous key that was provided with the data that identified the same individuals across datasets.

The Ministry of Health and Social Protection’s healthcare service database, known as the *Registro Individual de Prestación de Servicios de Salud* (RIPS), contains key information on health services provided including service provider information, diagnoses and treatments. RIPS data was similarly linked to the RUV and PAPSIVI service data.

### Measures

#### Outcome variables

PAPSIVI services are only open to registered victims of the armed conflict and attendance is recorded through the presentation of the Colombian national ID card, allowing attendance to be linked to registered victim status with the data provided to us only with pseudonymous identifiers. We included i) type of PAPSIVI session offered and ii) number of PAPSIVI sessions as outcome variables.

PAPSIVI offers psychosocial support sessions in either an individual, family, group or community setting. PAPSIVI employs a matched care model where intensity of intervention was matched to level of need. An initial assessment identifies the needs of the victim after which the most appropriate psychosocial care service is advised. If the victim’s needs exceed the scope of psychosocial care within the PAPSIVI, both in terms of severity and the type of care required, they are referred to specialised services outside of PAPSIVI. A brief outline of the sessions offered by PAPSIVI are outlined below.

<u>Individual sessions</u> – one-to-one psychosocial intervention for individuals considered to be severely impacted by the armed conflict. The recommended number of individual sessions is set at 8; however, if deemed necessary, additional sessions can be provided. Individual sessions are specifically recommended for victims of torture, the effects of sexual violence, forced disappearance, and forced recruitment to armed groups [12,13].

<u>Group sessions</u> – group sessions are offered as a variant of individual sessions and provide support to victims who have experienced similar victimising events or require support on a specific issue. There is a minimum of 5 and a maximum of 12 participants per group. Guidelines suggest 8 sessions [12,13].

<u>Family sessions</u> – psychosocial care for family groups where one or more family members have been victims of the armed conflict resulting in harm and emotional suffering to the family dynamic with a recommended number of 8 weekly sessions.

<u>Community sessions</u> – community psychosocial care refers to services offered in a collective setting and aims to repair damage to the social fabric of communities caused by the armed conflict. Support sessions are classified into 4 categories with a total of 17 subjects as sub-categories. For each person, 6 sessions are recommended. However, if a topic has been sufficiently worked on in fewer sessions another topic could be included in the remaining sessions in consultation with the community.

#### Exposure variables

Type of exposure to the armed conflict was taken from the victims’ registry (RUV) and included the following officially recognised categories: witness to terrorism or combat, sexual violence, forced disappearance, forced displacement, homicide, exposure to mines / improvised explosives, kidnapping, torture, child recruitment to armed groups, forced land abandonment / dispossession, loss of personal belongings, physical injuries, psychological injuries, threats, and confinement). Classification of the type of exposure is conducted during registration through interview and documentation.

Any previous psychiatric diagnoses were obtained through the RIPS database of healthcare contacts. These diagnoses were made independently of the PAPSIVI programme based on individuals’ contact with inpatient or outpatient health services and were recorded using the ICD-10 classification system. Diagnoses obtained after the individual’s first contact with the PAPSIVI service were not considered.

#### Covariates

We included socio-demographic variables (sex, ethnicity, age) as potential confounders in regression analyses. Age was considered as a continuous variable whilst ethnicity was categorised as minority ethnic group (Indigenous, Roma, Raizal, Afro-Colombian, Palenquero de San Basilio) and White/Mestiza. As no direct measure of socioeconomic status was available, we used healthcare regime as a proxy variable. Healthcare regime is a mixed public-private model in Colombia with a subsidised option for those eligible. Eligibility is calculated based on a poverty index that considers socioeconomic, demographic and welfare factors. Healthcare regime was recorded as either subsidised or contributory.

We also included a measure of mean conflict intensity at the municipality of registration, based on the Centre for Resource for Analysis of Armed Conflict (CERAC) database. CERAC’s conflict intensity measure is based on the presence of armed actors by year and the number of conflict events at the municipality level in Colombia and categorised into high or low intensity compared to the national average.

### Statistical analysis

Analysis was completed using *R* version 4.4.1 [22] on a Windows x86_64 secure analysis platform. All analysis code is available on the online archive: https://github.com/vaughanbell/papsivi_treat

Descriptive statistics were generated as frequencies and means with standard deviations and proportions as appropriate. Logistic regression models assessed the odds of attending each session type (individual, community, family, group) based on exposures. Cluster robust standard errors were calculated to account for clustering at the individual level using R package *sandwich*. This adjustment accounts for within-person correlation and dependencies between repeated observations for the same person. Linear regression models were used to assess associations between i) victimisation and ii) previous mental health diagnosis and number of PAPSIVI sessions by type. Cluster robust standard errors were similarly calculated. We conducted additional linear regression analyses using sex, ethnicity, and healthcare regime as exposure variables to explore their associations with number of PAPSIVI sessions.

#### Secondary analysis

We investigated whether associations between previous mental health and access to PAPSIVI differed by ICD diagnosis.

## Results

The data used by this study covers 2013-2021 and includes 8,359,538 individuals who registered as victims with the RUV, of whom 534,818 (6.4%) accessed PAPSIVI services for conflict victims. Table 1 displays demographic and descriptive statistics. In total, 502,814 (94.0%) individuals attended only one type of PAPSIVI session whilst 32,004 (6.0%) attended more than one type.

**Table 1.** Demographic and descriptives of all registered victims and PAPSIVI service users. PAPSIVI = Comprehensive Psychosocial Care and Health Program for Victims (Programa de Atención Psicosocial y Salud Integral a Víctimas). RUV = Central Registry for Victims (Registro Unico de Víctimas).

|  | *All RUV registered victims* |  | *Accessed PAPSIVI* |  |
| --- | --- | --- | --- | --- |
|  | *Total N = 8,359,538* |  | *Total N = 534,818* |  |
|  | *Mean* | *SD* | *Mean* | *SD* |
| *Age* | 34.7 | 20.6 | 41.2 | 19.1 |
| Missing | 25217 | 0.3% | 140 | <0.01% |
|  | *N* | *%* | *N* | *%* |
| *Sex* |  |  |  |  |
| Male | 4,240,400 | 50.7% | 201,666 | 37.7% |
| Female | 4,114,735 | 49.2% | 332,811 | 62.2% |
| Other | 3,856 | <0.01% | 321 | 0.1% |
| Missing | 547 | <0.01% | 20 | <0.01% |
| *Ethnicity* |  |  |  |  |
| Indigenous | 1,061,174 | 12.7% | 96,282 | 18.0% |
| Roma | 223,058 | 2.7% | 17,776 | 3.3% |
| Raizal | 7,280 | 0.1% | 469 | 0.1% |
| Afro-Colombian | 10,426 | 0.1% | 827 | 0.2% |
| Palenquero de San Basilio | 8,885 | 0.1% | 415 | 0.1% |
| White/Mestiza | 7,048,715 | 84.3% | 419,049 | 78.4% |
| *Healthcare regime* |  |  |  |  |
| Contributory | 1,987,877 | 23.8% | 100,394 | 18.8% |
| Subsidised | 5,088,138 | 60.9% | 402,431 | 75.2% |
| Missing | 1,283,523 | 15.4% | 31,993 | 6.0% |
| *Disability* |  |  |  |  |
| No | 7,967,800 | 95.3% | 495,374 | 92.6% |
| Yes | 391,738 | 4.7% | 39,444 | 7.4% |
| *Previous psychiatric diagnosis* |  |  |  |  |
| No | 7,888,105 | 94.36% | 510,906 | 95.53% |
| Yes | 471,433 | 5.64% | 23,912 | 4.47% |

Of the 534,818 people who accessed PAPSIVI, 23,912 had a mental health diagnosis pre-contact with PAPSIVI. Supplementary Table 1 displays diagnosis broken down by ICD categories. Table 2 displays the breakdown of PAPSIVI services attended by session type.

**Table 2.** Number of attendees by total number of sessions attended in each intervention modality. Note: some individuals accessed more than one type of modality.

| *Total*<br>*sessions*<br>*attended* | *Intervention type* |  |  |  |  |  |  |  |
| --- | --- | --- | --- | --- | --- | --- | --- | --- |
|  | *Individual* |  | *Group* |  | *Family* |  | *Community* |  |
|  | *N* | *%* | *N* | *%* | *N* | *%* | *N* | *%* |
| 1 | 37,918 | 28.7% | 130 | 2.8% | 5,785 | 1.9% | 1,972 | 1.5% |
| 2 | 11,449 | 8.7% | 58 | 1.3% | 4,938 | 1.6% | 1,809 | 1.4% |
| 3 | 9,442 | 7.2% | 34 | 0.7% | 3,957 | 1.3% | 2,230 | 1.7% |
| 4 | 3,399 | 2.6% | 73 | 1.6% | 4,161 | 1.4% | 7,114 | 5.5% |
| 5 | 4,902 | 3.7% | 84 | 1.8% | 9,526 | 3.1% | 20,241 | 15.6% |
| 6 | 7,765 | 5.9% | 137 | 3.0% | 25,132 | 8.3% | 80,124 | 61.9% |
| 7 | 13,969 | 10.6% | 320 | 7.0% | 53,847 | 17.8% | 9,798 | 7.6% |
| 8 | 35,901 | 27.2% | 899 | 19.7% | 179,729 | 59.4% | 3,019 | 2.3% |
| 9 | 4,176 | 3.2% | 2,733 | 59.9% | 10,197 | 3.4% | 434 | 0.3% |
| 10 | 1,633 | 1.2% | 82 | 1.8% | 2,121 | 0.7% | 726 | 0.6% |
| 11 | 572 | 0.4% | 20 | 0.4% | 641 | 0.2% | 381 | 0.3% |
| 12 | 317 | 0.2% | 2 | 0.0% | 458 | 0.2% | 845 | 0.7% |
| ≥13 | 604 | 0.5% | 2 | 0.0% | 2,112 | 0.7% | 652 | 0.5% |
| Total | 132,047 |  | 4,564 |  | 302,604 |  | 129,381 |  |

The distribution of attended sessions peaks at, or near, the recommended number of sessions for interventions (8 for individual, group and family sessions; 6 for community sessions). Although the majority of participants who received individual sessions received multiple sessions, the single most frequent number of individual sessions received is 1, equivalent to 28.71% of attendees to individual sessions receiving a single session.

### Intervention provision

As displayed in Figures 1a – 1d, we found varying associations between victimisation exposure and type of PAPSIVI sessions provided in models adjusted for age, ethnicity, sex, healthcare regime and CERAC.

**Figure 1a.**
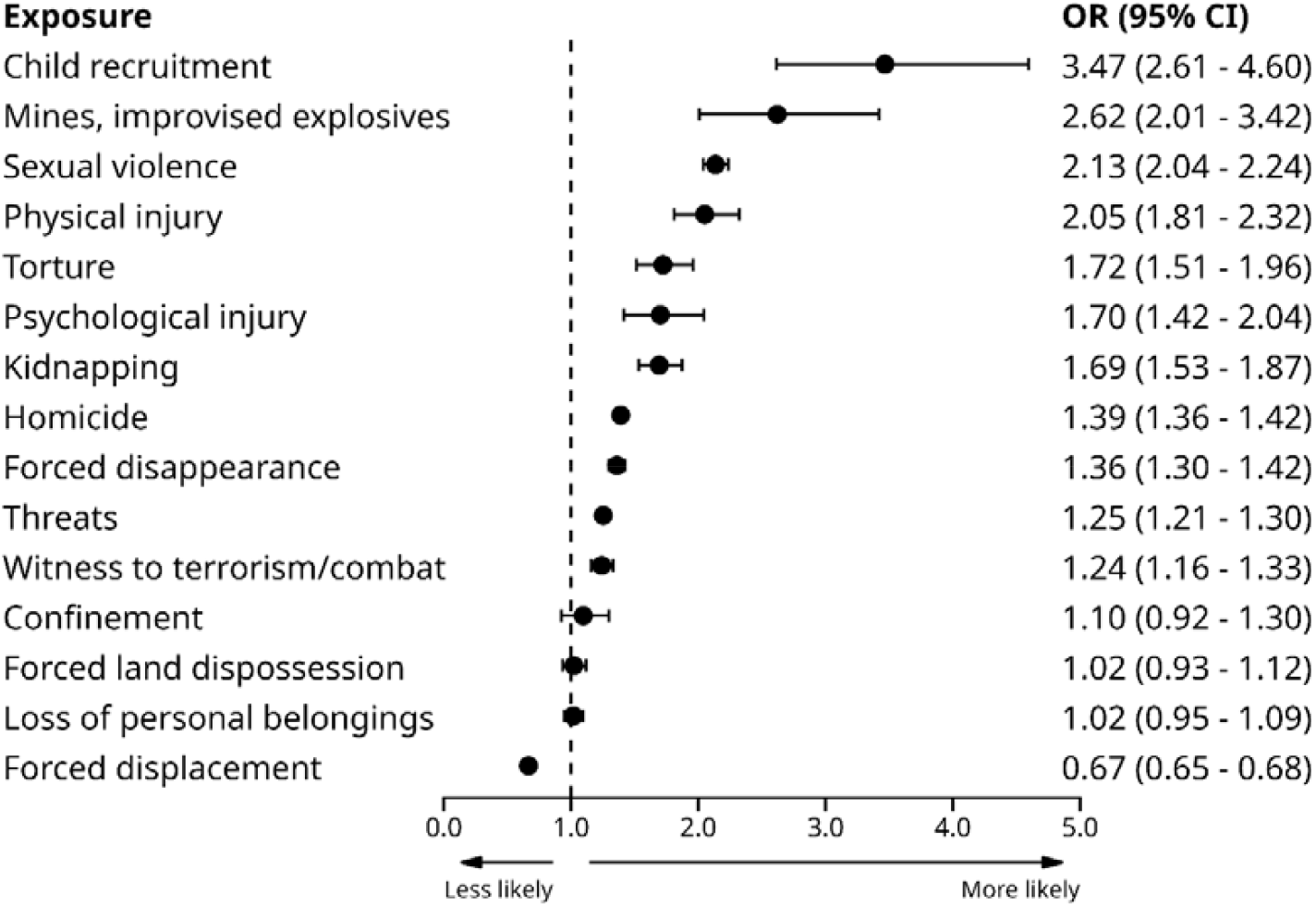
Odds of attendee receiving individual sessions by armed conflict exposure type

**Figure 1b.**
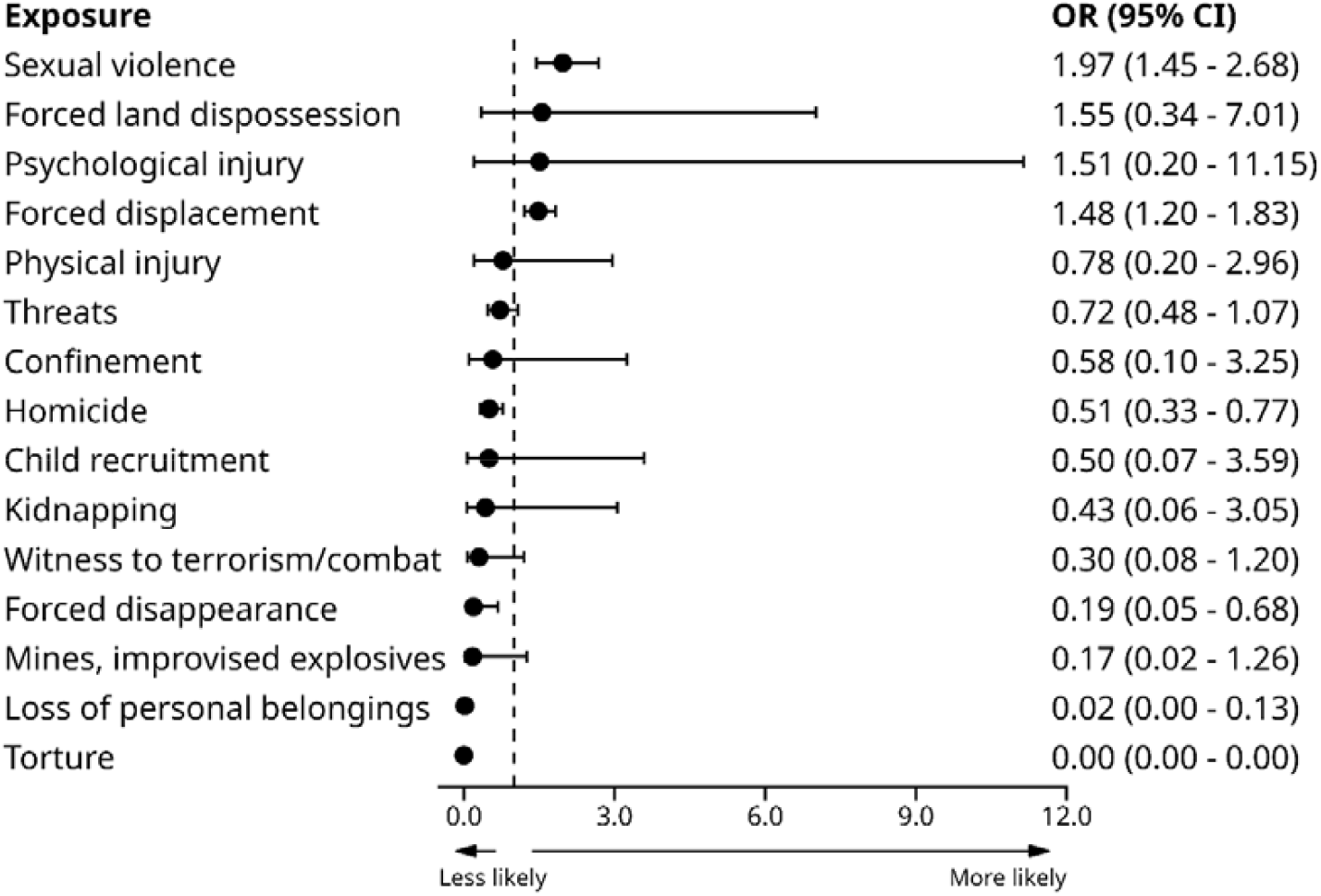
Odds of attendee receiving group sessions by armed conflict exposure type

**Figure 1c.**
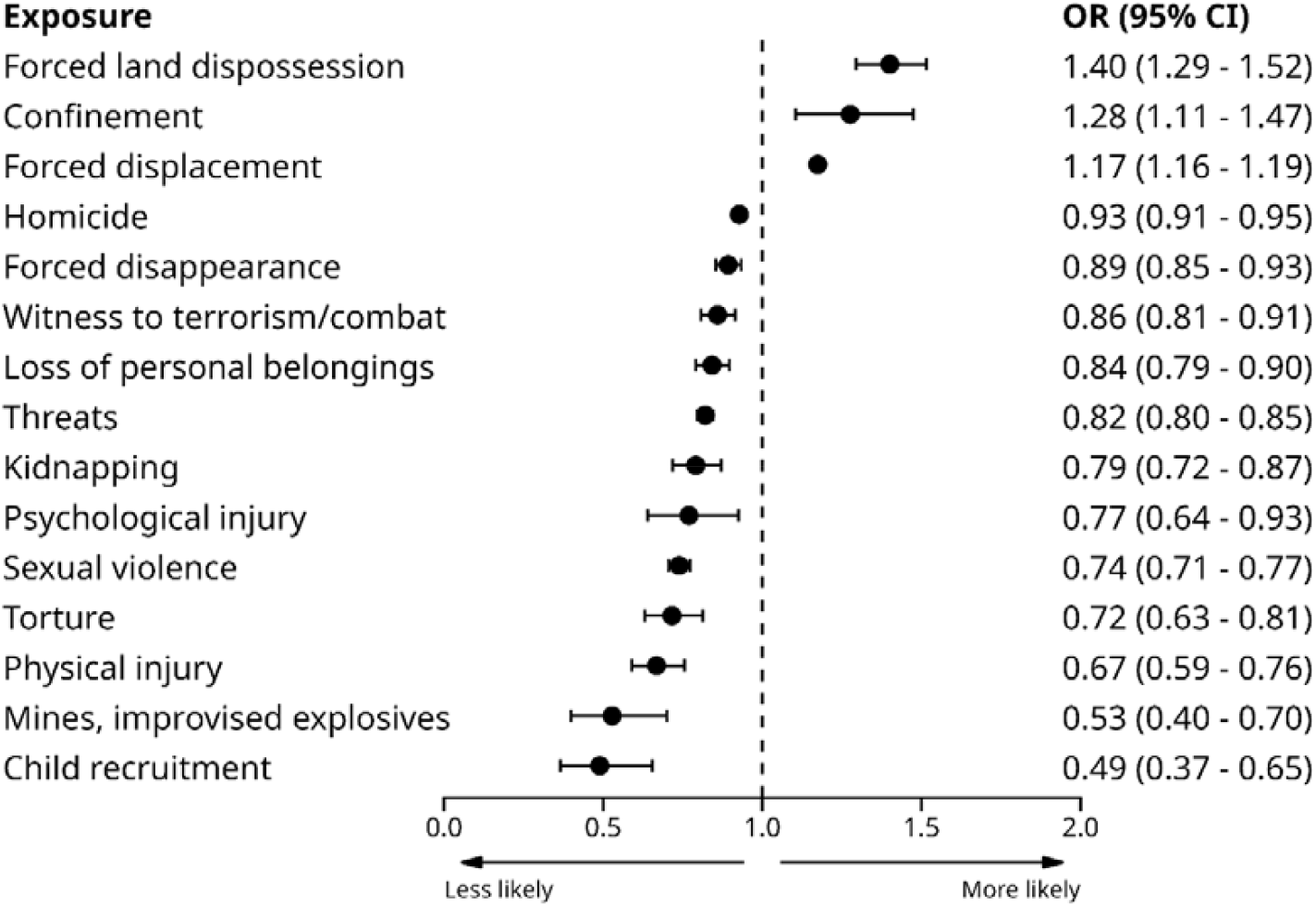
Odds of attendee receiving family sessions by armed conflict exposure type

**Figure 1d.**
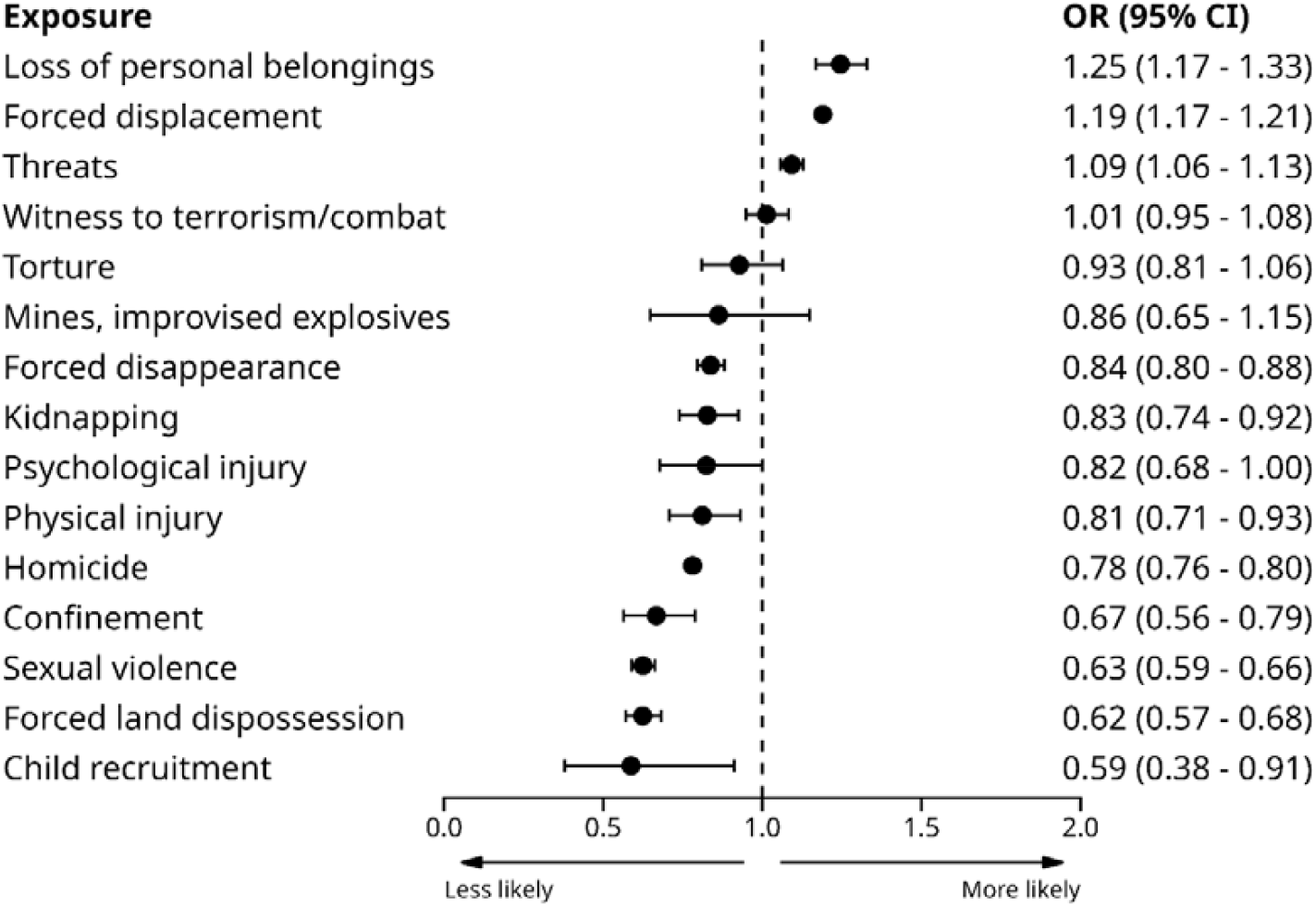
Odds of attendee receiving community sessions by armed conflict exposure type

For individual sessions, results show prioritisation largely in line with PAPSIVI guidelines where exposure to torture, sexual violence, forced disappearance, and child recruitment were associated with higher levels of access to individual sessions. More broadly, exposures that reflect individual affectation are more likely to receive individual sessions, whereas those reflecting group impact are more likely to receive group, family or community sessions. Notably, estimates for group-based sessions had large and uneven confidence intervals due to the low overall number of such sessions, resulting in imprecise estimates for some exposure types. The notable exception was sexual violence, which showed greater odds of both individual and group sessions.

Attendees with any prior mental health diagnosis had increased odds of attending either individual sessions (OR = 1.24; 95% CIs 1.18; 1.30) or family sessions (OR = 1.30; 95% CIs 1.25; 1.35) but lower odds of attending either group sessions (OR = 0.48; 95% CIs 0.37; 0.63) or community sessions (OR = 0.53; 95% CIs 0.50; 0.55), broadly in line with PAPSIVI recommendations for the use of more individualised interventions for those with higher psychosocial needs. There were few associations between specific diagnostic group and session type (see Supplementary Table 2), likely reflecting the low rate of prior diagnosis, although diagnoses outside the scope of PAPSIVI tended to have lower rates of individual session provision (e.g. behavioural syndromes associated with physiological disturbances, F50-F59; childhood onset disorders, F90-98; developmental disorders, F80-89; and intellectual disabilities, F70-79).

### Intervention engagement

Table 3 shows the results of adjusted regression analyses indicating the relative number of sessions by exposure type. Those with exposure to physical injury typically received the highest number of sessions across modalities, followed by exposure to torture, mines / improvised explosives, land dispossession and confinement. We note those with exposure to displacement tended to attend a relatively lower numbers of sessions.

**Table 3.** Adjusted associations between victimisation and number of sessions of PAPSIVI attended by type using mean difference of number of sessions between victimisation and non-victimisation.

| *Victimisation* | *Number of sessions mean difference (95% CI)* |  |  |  |
| --- | --- | --- | --- | --- |
|  | *Community* | *Family* | *Group* | *Individual* |
| Torture | 11.4 (9.2, 13.5) | 15.2 (12.3, 18.1) | - | 13.5 (9.7, 17.3) |
| Sexual Violence | 8.6 (7.6, 9.5) | 9.4 (8.71, 10.2) | 5.9 (0.8, 11.0) | 9.0 (8.2, 9.8) |
| Threats | 7.3 (6.9, 7.6) | 9.7 (9.3, 10.0) | 10.6 (3.5, 17.7) | 8.6 (8.1, 9.2) |
| Displacement | -7.2 (-7.4, -7.1) | -8.5 (-8.7, -8.4) | -7.8 (-11.1, -4.4) | -5.5 (-5.7, -5.3) |
| Homicide | 4.9 (4.6, 5.1) | 4.7 (4.5, 4.9) | 3.5 (-1.3, 8.2) | 1.5 (1.2, 1.7) |
| Witness to terrorism | 8.7 (7.8, 9.6) | 12.1 (11.1, 13.2) | - | 11.1 (9.7, 12.4) |
| Physical injury | 14.3 (11.7, 16.9) | 17.6 (15.7, 19.6) | 4.6 (4.1, 5.1) | 17.4 (14.1, 20.8) |
| Land dispossession | 9.2 (7.9, 10.6) | 13.3 (12.3, 14.4) | 13.7 (13.2, 14.2) | 12.4 (10.8, 14.1) |
| Confinement | 8.5 (6.0, 11.0) | 14.7 (12.5, 16.8) | - | 16.9 (11.7, 22.2) |
| Psychological injury | 7.8 (5.6, 9.9) | 10.5 (7.8, 13.2) | 24.8 (23.6, 26.0) | 8.2 (4.9, 11.5) |
| Mines, explosives | 10.0 (5.8, 14.2) | 11.6 (7.6, 15.6) | -5.6 (-5.8, -5.3) | 12.9 (8.1, 17.7) |
| Loss of belongings | 8.4 (7.7, 9.1) | 11.8 (11.0, 12.6) | - | 8.2 (7.1, 9.3) |
| Kidnapping | 5.7 (4.2, 7.1) | 10.3 (8.8, 11.8) | - | 6.7 (5.3, 8.2) |
| Child recruitment | 2.3 (-1.5, 6.2) | 10.8 (6.3, 15.3) | - | 3.7 (0.2, 7.1) |
| Disappearance | 5.1 (4.5, 5.7) | 5.0 (4.6, 5.5) | -0.8 (-1.2, -0.4) | 2.6 (2.0, 3.2) |

For key marginalised groups, when adjusted for covariates, female sex was associated with attending more individual sessions (beta = 1.03; 95% CIs 0.92; 1.14), family sessions (beta = 0.64; 95% CIs 0.58; 0.70), community sessions (beta = 0.43; 95% CIs 0.34; 0.52) but not group sessions (beta = 0.20; 95% CIs -0.11; 0.52). Ethnic minority status was associated with attending a greater number of individual sessions (beta = 0.44; 95% CIs 0.31; 0.58), family sessions (beta = 0.60; 95% CIs 0.52; 0.69), and community sessions (beta = 0.43; 95% CIs 0.33; 0.53). Ethnic minority status was not reliably associated with number of group sessions received as the confidence intervals of the estimate crossed zero (beta = -0.51; 95% CIs - 1.02; 0.00). Being on a subsidised healthcare regime was not associated with reliable differences in attending group sessions (beta = -0.15; 95% CIs -0.74; 0.44), family sessions (beta = 0.04; 95% CIs -0.04; 0.11) and community sessions (beta = 0.04; 95% CIs -0.08; 0.15) but was reliably associated with attending a lower number of individual sessions (beta = -0.42; 95% CIs -0.56; -0.28).

Previous mental health diagnosis was associated with a substantially greater number of individual sessions attended (beta = 4.30; 95% CIs 3.94; 4.66), slightly greater numbers of family (beta = 0.91; 95% CIs 0.73; 1.09) and community sessions (beta = 0.45; 95% CIs 0.18; 0.72), and no reliable difference in the number of group sessions (beta = 1.23; 95% CIs -0.19; 2.66).

## Discussion

In this national study of over half a million users of the PAPSIVI psychosocial support service for victims of the Colombian armed conflict, we examined to what extent conflict victimisation was associated with intervention provision and engagement. We additionally examined associations between these metrics of service quality and prior diagnosis of psychiatric disorder, given that pre-existing mental health conditions should be an indicator of psychosocial need. The results showed that patterns of intervention provision and engagement were broadly in line with PAPSIVI guidance, with victims with conflict exposures reflecting higher levels of individual affectation being more likely to be provided individual intervention sessions and more likely to receive a greater number of sessions. Female sex, ethnic minority status, and being on a subsidised healthcare regime were typically associated with attending a greater number of sessions, indicating higher levels of engagement in priority groups.

The results indicate that patterns of provision and engagement generally align with PAPSIVI guidelines indicating that PAPSIVI is broadly meeting its criteria for service provision by providing recommended intervention modalities by conflict exposure type and maintaining engagement for those who attend. We would not necessarily expect perfect alignment with recommendations and interventions offered for specific exposures because practitioners may offer interventions on the specific features of the individual which may not be fully captured by a single classification of a conflict exposure. Furthermore, formal recommendations were only published in 2018 which may have led to variable alignment before this date, and some exposures (for example, sexual violence) may have been under-reported at victim registration but may come to light during the assessment process with trained practitioners. We also note that more severe armed conflict exposures may be more likely to co-occur with other exposures, potentially explaining why exposure to sexual violence is associated with an increased number of sessions across modalities, despite recommendations specifically suggesting individual and family sessions as preferred intervention modalities. Nevertheless, there is clear evidence that recommendations have an important influence on both provision and engagement, indicating a strong influence of policy on practice.

However, we note some findings which may indicate areas of improvement. For individual sessions, although the majority of attendees received multiple sessions and there was a second peak at the recommended number of sessions (eight), the most frequent number of attended sessions was one. It is possible there are several explanations for single-session attendees, including therapists deciding to refer to a more appropriate intervention modality after commencement of sessions, either inside or outside of PAPSIVI, or attendees not being able to remain for future sessions due to displacement or migration. However, the fact that almost 29% of individual session attendees receive only one session may also reflect an influence of early drop-out in a course of intervention.

Dropout rates for psychological interventions are typically 20%, a figure mostly drawn from trials in Europe and the United States [23,24]. Dropout in an intervention study of individuals exposed to the Colombian armed conflict was reported as 36% [25]. A study of dropout from mental health treatment across six countries in the Americas (Argentina, Brazil, Colombia, Mexico, Peru and USA) identified the influence of both structural and attitudinal barriers on non-attendance [26]. Both of these have been widely documented as affecting healthcare access for victims of the Colombian armed conflict where economic barriers [27,28], hard-to-navigate bureaucracy [27], lack of sensitivity to local context [29] and stigma [30,31] have been identified as significant obstacles to engagement.

The results from this study and a previous study on access to PAPSIVI [9] show that key marginalised groups – women, ethnic minorities, and those on state subsidised healthcare insurance schemes – typically have higher levels of access to PAPSIVI and its programmes. Nevertheless, it remains unclear to what extent these promising results are meeting the extent of needs in these key populations. A survey of professionals working in PAPSIVI reported service implementation barriers as well as perceived difficulties in delivering interventions that were culturally and ethnically appropriate [32]. Qualitative studies have reported concerns that some PAPSIVI interventions are poorly suited to local contexts [33] and may compound discrimination [34]. Although the generalisability of these findings to PAPSIVI as a whole is uncertain, they highlight the need for an in-depth analysis of the structural, cultural and social barriers to access and engagement to improve service delivery.

We also note an important feature of PAPSIVI service delivery, in that group sessions were infrequently used, consisting of only 0.8% of the total sessions delivered. This accounts for why it was not possible to calculate estimates for associations between some exposures and group sessions (Table 3) and why there are large confidence intervals in Figure 1b, as there were few instances on which to base estimates. However, this may also reflect the fact that group sessions are not meeting the needs of the service / and or attendees and may be better covered by other intervention modalities.

We note several limitations with this study. Data were from service records and it was not possible to verify the accuracy of data recording. Principally, conflict exposure data was from the national register of victims which requires only one verified conflict exposure for registration, which means this data likely undercounts total conflict exposure when individuals have multiple affectations. In addition, the available data lacked direct measurements of socioeconomic status. The binary distinction between subsidised and contributory healthcare insurance regime was used as a proxy. Although this classification is based on an official poverty index, it lacks the representation of the full range of socioeconomic variation.

Importantly, this study evaluated quality of service delivery using routine service data but not effectiveness of the service in ameliorating psychosocial harms – its main aim. PAPSIVI does not routinely collect specific outcomes on effectiveness of the interventions. A previous study on 771 PAPSIVI attendees suggested high levels of satisfaction with the service [21] but given that the service is national, and therefore serves diverse populations across Colombia who have different needs at the levels of both conflict exposure and community, it is likely that there are important predictors of service quality and effectiveness that are not captured by the current study.

## Conclusions

The present study offers one of the most comprehensive evaluations of nation-wide mental health services for conflict-affected victims. We report that intervention modalities are generally provided in line with PAPSIVI guidelines and that key priority groups – such as women, ethnic minorities, and those on subsidised healthcare – are more likely to engage with services. However, further work is needed to understanding factors driving service disengagement in those who do not receive adequate support, and effectiveness of the interventions.

## Data Availability

The data used in this study was requested after application to the Colombian Ministry of Health and has not been made publicly available due to that agreement. All analysis code is available online at https://github.com/vaughanbell/papsivi_treat

## Declarations

### Consent for publication

This manuscript does not contain any individual person’s data in any form.

### Availability of data and materials

The data used in this study was requested after application to the Colombian Ministry of Health and has not been made publicly available due to that agreement. All analysis code is available online at https://github.comvaughanbell/papsivi_treat

### Authors’ contributions

VB, FI, RS and FS conceptualised the study. All authors were involved in the consultation on study design and interpretation of the findings. CCF and VB developed the analysis code. CCF and VB drafted the initial manuscript with input from all authors.

## Acknowledgements

Many thanks to Mariana Matamoros Cárdenas for her assistance with data preparation.

